# Healthy diet perceptions and drivers of fruit and vegetable food choices among adolescents in Benin: a qualitative study

**DOI:** 10.64898/2026.06.30.26356853

**Authors:** Ampa Dogui Diatta, Fifali Sam Ulrich Bodjrènou, Abdoulaye Pedehombga, Sarah Eissler, Irene Medeme Mitchodigni, Kenda Cunningham, Deanna Olney, Lilia Bliznashka, Elyse Iruhiriye

## Abstract

Globally, 75% of adolescents do not meet recommendations for fruits and vegetables (F&V) consumption. This study investigated drivers of F&V choices among adolescents in Benin and identified barriers and facilitators to modifying F&V consumption. We conducted 16 focus group discussions (FGDs) with 126 adolescents and 5 semi-structured school observations in December 2024. FGDs were purposively chosen to explore variation by region (north/south), school location (urban/rural), adolescent age (12–15 years/16-18 years) and gender (boys/girls). Inductive and deductive thematic content analysis was performed using *Atlas.ti*. Findings showed that adolescents perceived a healthy diet as one composed of meals providing nutrients and strength, and including F&V. At home, adolescents mentioned eating staples and other vegetables more than other food groups. The foods adolescents reported typically eating at school varied by age, gender, and location. The primary categories of factors that influenced adolescent F&V choices were: intrapersonal (knowledge related to healthy eating and the nutrition and health benefits of F&V, taste preferences, and health prioritisation), socio-cultural (family and peer influences), and food environment (low availability, low affordability, convenience, and desirability). DFC factors were consistent across adolescent age, gender, and location. Multiple, dynamic, and multilevel factors influence adolescent F&V choices. Interventions that simultaneously address multiple barriers and involve family and peers are likely to be more successful in promoting F&V consumption and healthy diets than interventions only addressing individual barriers.

**Key messages:**

- According to adolescents in Benin, a healthy diet was one composed of meals providing nutrients and strength and including fruits and vegetables (F&V).
- Intrapersonal, socio-cultural, and food environment factors influenced adolescent F&V choices regardless of adolescent gender, age, and location.
- Adolescent F&V choices are influenced by a dynamic interplay of multiple factors. Thus, interventions to improve adolescent diets should likely address several critical barriers in an integrated and holistic manner.

## 1. Introduction

Inadequate diets, including low fruit and vegetable (F&V) consumption, increase preventable morbidity and mortality from non-communicable diseases (NCDs) (Afshin et al., 2019; Miller et al., 2019; Yip et al., 2019). In 2023, dietary risk factors contributed to over 7 million deaths and 186 million disability-adjusted life-years globally (IHME, 2026). In Benin, dietary risk factors have remained the fifth highest ranking risk factor for disease since 1990 (IHME, 2015).

Adolescence is a critical life stage of developing taste preferences, growing autonomy in food choices, and increasing peer and food environment (FE) influences (Neufeld et al., 2022). Eating patterns and habits established during adolescence persist later in life and affect health outcomes both in adolescence and adulthood (Neufeld et al., 2022; Twig et al., 2016; Zheng et al., 2025). Additionally, nearly one-half of adolescents globally are experiencing four or more NCD risk factors (Biswas et al., 2022). This trend has resulted in increasing NCD prevalence among adolescents, including overweight/obesity and hypertension (Guo et al., 2026; Zhou et al., 2026).

In Benin, 12% of adolescents 13–17 years of age were living with overweight or obesity in 2016, with higher prevalence among girls than boys (19% vs 8%) (WHO, 2016). Like other low- and middle-income countries, Benin is undergoing a nutrition transition marked by changing FE and increased intake of fats and sugars among urban residents (Delisle et al., 2012; Sodjinou et al., 2009). Simultaneously, diets remain monotonous with low F&V consumption across all population groups (Bliznashka et al., 2024). More than one-half of Benin’s food supply comes from cereals, roots, and tubers, with insufficient F&V supply to meet dietary recommendations (Our World in Data, 2021). This low F&V availability contributes to the high proportion (81%) of adolescents not meeting global or national recommendations for F&V consumption (PNLMNT, 2011).

Despite their importance, adolescent diets and the factors that influence them are poorly understood in Benin. A recent scoping review identified only six articles that examined F&V consumption in adolescents, all relying on data collected more than a decade ago (Bliznashka et al., 2024). Existing research on adolescent drivers of food choice (DFC) is also limited. One study, conducted 15 years ago, showed that low F&V availability and accessibility, limited health and nutrition knowledge, higher availability and accessibility of unhealthy foods, and food safety concerns hindered F&V consumption (Nago et al., 2012). A more recent study conducted in 2020 found that limited dietary knowledge, low affordability of nutrient-dense foods, and peer pressure explained unhealthy food choices among adolescent girls (Mama Chabi et al., 2022). Nevertheless, both studies were conducted in Cotonou, the largest urban centre in Benin, providing limited understanding of how these and other DFC factors affect adolescents’ food choices in rural settings and how F&V-related DFC have changed over time.

Understanding the perceptions and factors that influence food choices during adolescence is crucial to provide an entry point for targeted and cost-effective interventions to improve diets and reduce adolescent malnutrition. This study used ethnographic and anthropological approaches to: (1) explore school-going adolescents’ food choices, (2) identify the drivers underpinning F&V choices and consumption, and (3) identify barriers and facilitators to modify F&V food choices and increase F&V consumption.

## 2. Methods

### 2.1 Conceptual framework

DFC are individual-based motives that underpin decision-making about foods; these are shaped by a complex interplay of psychological, economic, social, cultural, environmental, and political factors (Blake et al., 2021; Karanja et al., 2022). In this study, we used a conceptual framework that captures different levels of influences (**Table 1**), adapted from previous literature (Boncyk, 2023; Karanja et al., 2022). This is important for adolescents who are both gaining food choice autonomy and relying on parents for most of their food intake (Karanja et al., 2022; Turner et al., 2020).

**Table 1.** Conceptual framework on the drivers of food choice^1^.

|  | Intrapersonal drivers | Socio-cultural drivers | Food environment drivers | Material assets and resources |
| --- | --- | --- | --- | --- |
| Factors | • Knowledge, attitudes, beliefs, and perceptions<br>• Preferences<br>• Goals and prioritisation<br>• Habits and routines<br>• Self-efficacy<br>• Identity<br>• Personality<br>• Mood<br>• Motivation<br>• Ethical concerns<br>• Physiological needs | • Social interactions<br>• Family influence<br>• Media influences<br>• Peer relationships<br>• Cultural values and practices<br>• Food tradition and customs<br>• Food taboos<br>• Religion<br>• Gender equity<br>• Women's empowerment<br>• Life course perspectives | • Availability<br>• Accessibility<br>• Affordability<br>• Convenience<br>• Desirability | • Income<br>• Food, water, and housing security<br>• Facilities |

<sup>1</sup> Adapted from Boncyk et al. 2023 and Karanja et al. 2022 (Boncyk, 2023; Karanja et al., 2022).

### 2.2 Study Design

This study was part of a larger qualitative study exploring the DFC with respect to F&V consumption among women of reproductive age (18-49 years of age) and adolescents (12-18 years of age) in Benin. Here, we present findings from focus group discussions (FGDs), free listing, and pile sorting with adolescents, and semi-structured observations (SSOs) at school.

The FGDs aimed to understand how adolescents perceive F&V food choices and why they adopt certain F&V eating behaviours. The free listing was designed to determine the salience of different factors on F&V food choices, whereas the pile sorting was designed to rank factors that promote or hinder F&V consumption. The SSOs were used to understand food availability in/around schools included in the study.

### 2.3 Study Setting

The study was conducted in the Atacora and Atlantique departments (north and south of the country, respectively), which have a high prevalence of food insecurity: 38.4% and 45.5%, respectively (WFP, 2026). Although recent data on adolescents are lacking, data from 2017-18 on women of reproductive age (15-49 years of age) indicate Atacora has a high prevalence of underweight (15.4%) and Atlantique has a high prevalence of overweight/obesity (31.4%) (INSAE & ICF, 2019).

### 2.4 Sampling

We used purposive sampling for both the schools and students in the study. The two departments were selected based on existing activities under a larger agri-food systems project (Olney et al., 2021) and to capture heterogeneity by agro-ecological characteristics, vulnerability to food insecurity, and malnutrition prevalence (Bodjrènou et al., 2023). In each department, communes (department sub-divisions) were stratified into urban/peri-urban or rural. One urban and one rural commune were purposively selected in each department based on the availability of large F&V markets in their catchment areas (**Figure 1**). In each commune, we purposively selected one middle school with adolescents aged 12-15 years and one high school with adolescents aged 16-18 years for a total of eight schools. All schools were public and mixed gender.

**Figure 1.**
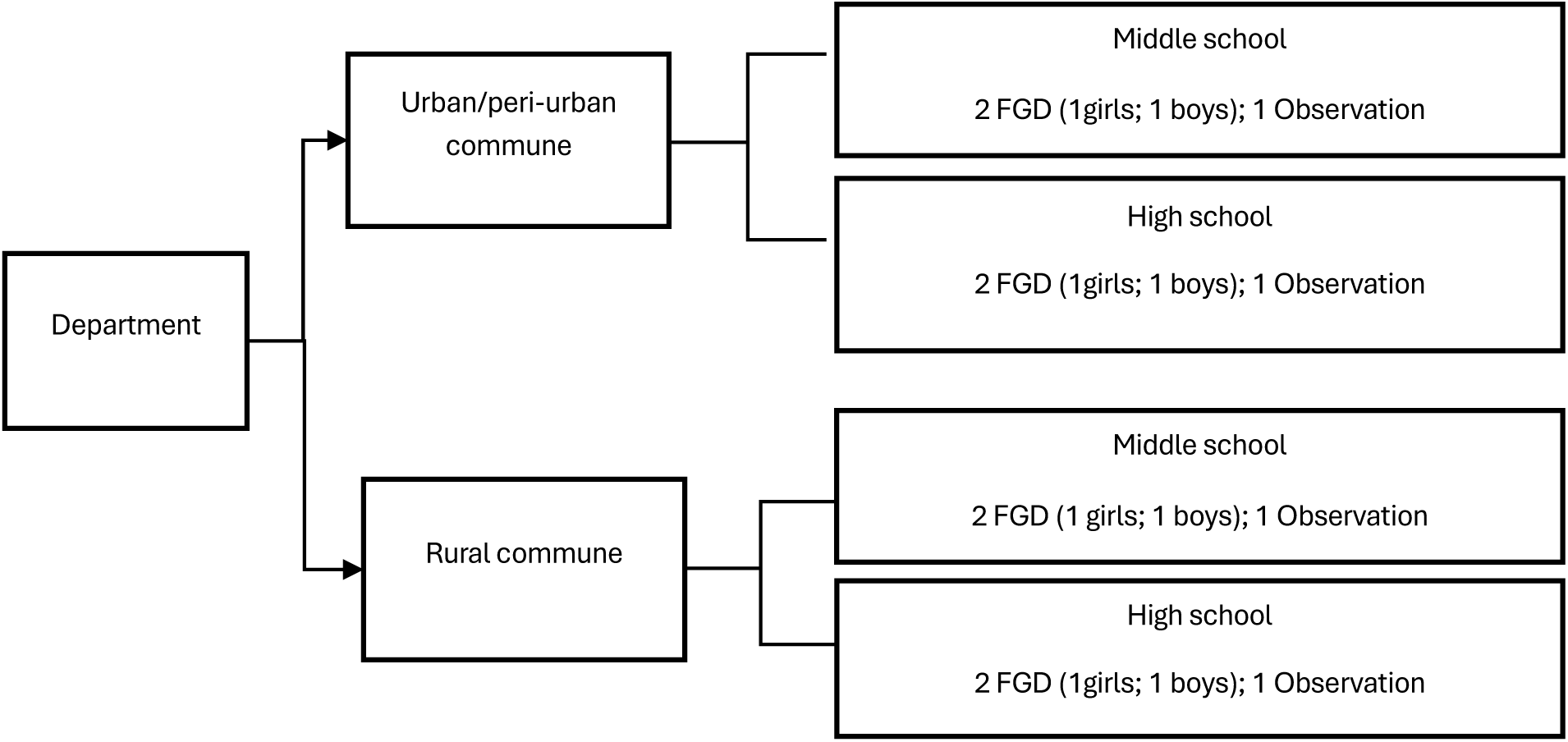
Sampling of schools included in the study. Abbreviations used: FGD, focus group discussion

In each school, two FGDs were conducted, one with girls and one with boys, followed by free listing and pile sorting exercises. The students who participated in the FGDs were purposively selected by the school principal based on their availability and willingness to participate. Each FGD had eight participants (four students per grade from two grade levels), except for one which had six participants. This sample of 16 FGDs with 126 participants was considered sufficient to reach saturation (Hennink & Kaiser, 2022). Five SSOs were conducted in and around the sampled schools. Eight SSOs were initially planned, but in some communes, the sampled schools were in the same building.

### 2.5 Data collection

The FGD, pile sorting, free listing, and SSO topic guides were developed in French by the study team. The FGD guide covered themes on food choices and practices (including gender differences), evolution of dietary patterns, perceptions about healthy diets, perceptions about F&V, barriers and facilitators to F&V consumption, and perceptions about information needs related to healthy diets and F&V consumption. Each FGD was followed by free listing and pile sorting exercises, which were primarily focused on F&V. FGDs lasted 87 minutes on average (range: 62-112 minutes). The free listing and pile sorting lasted 52 minutes on average (range: 23-82 minutes).

The SSO guide included a checklist of foods available in/around the school. All vendors located within the school or within a 100-meter radius around the school and open on the day of the observation were covered. The SSO guide also included questions about school meals. None of the schools included in our sample had a school meal program at the time of the study. SSOs were held during three morning and two lunch breaks. The SSOs covered all breaks for their entire duration with the morning breaks lasting 20-30 minutes and the lunch breaks lasting 1-1.5 hours.

Data were collected in December 2024 by six Beninese research assistants (RAs) (5 female and 1 male), all of whom had at least a master’s degree in nutrition, sociology, or anthropology and extensive experience in qualitative data collection. They were fluent in French and at least one local language. FDGs, pile sorting, and free listing were conducted in French, mixed with a local language if needed to provide further explanations. Each FGD was facilitated by two RAs, a moderator and a notetaker.

RAs participated in a five-day training provided by the study team, which covered research ethics, qualitative data collection methods, and detailed reviews of the topic guides. The FGD and SSO guides were pilot tested in a non-study area, and refinements were made based on the pilot.

### 2.6 Data Analysis

RAs audio-recorded and transcribed FGDs verbatim in French. Data from FGDs were analysed in *Atlas.ti Web* using thematic content analysis. We used a hybrid approach of deductive and inductive coding. Deductive codes were based on the conceptual framework described in section 2.1. Inductive codes emerged during data analysis, and reflected broader, context-specific themes. An initial codebook was developed and refined during double coding of two transcripts. The remaining transcripts were coded by one researcher and reviewed by another. Regular debriefings were conducted between the two researchers to ensure coding consistency and discuss emergent findings. Example quotes for each theme were extracted and translated to English.

Data from the free listing, pile sorting, and SSOs were analysed in Excel. For free listing, facilitators and barriers to F&V consumption were analysed inductively, with themes mapped to the conceptual framework. For each factor, we counted the number of mentions in total, and by region, location, and age. For pile sorting, a total of 15 factors were mentioned across all FGDs. For each FGD, factors were ordered from most to least important and assigned the rank of that factor. For example, if 15 factors were reported in an FGD, the highest ranking or most important one was assigned a score of 15. For SSOs, we reported food availability at the food group level. A food group was considered available if at least one vendor sold at least one item in that food group. For all analyses, foods were grouped according to the FAO guidelines for constructing the minimum dietary diversity indicator: (1) grains/roots/tubers; (2) pulses; (3) nuts/seeds; (4) dairy; (5) meat/poultry/fish; (6) eggs; (7) dark green leafy vegetables; (8) other vitamin A–rich fruits and vegetables; (9) other vegetables; and (10) other fruit (FAO, 2021).

Findings from the FGDs, free listing, pile sorting, and SSOs were triangulated by themes to synthesise results. Although the study focused on F&V, animal source foods (ASF) emerged prominently in the results. We, therefore, highlight findings on ASF throughout the results and discussion, even though they were not explicitly included in the study design. A community validation workshop was organized in October 2025 to share results with study participants and to inform the interpretation of results.

### 2.7 Research Team and Reflexivity

The study was implemented by a team of national and international researchers, all with prior experience conducting qualitative studies and DFC studies in Africa. Two team members are from Benin and two are from other West African countries. Data were analysed by a female American and a male Senegalese, both native French speakers. Biases during analysis were mitigated by iterative discussions with the wider team. Results were also validated by the study participants during the community validation workshop.

### 2.8 Ethical Considerations

Ethical clearance was obtained from the Comité d’Ethique de la Recherche de l’Institut des Sciences Biomédicales Appliquées (CER-ISBA), Cotonou-Bénin (N°222) and from the Institutional Review Board of the International Food Policy Research Institute (NDH-24-1144). Written permissions were also obtained from the Ministry of Education, local authorities, and the principals of the schools where the study was conducted. RAs provided information about the study in French. Written assent was obtained from all adolescents and written consent from the school principal. Students received a snack as appreciation for their time.

## 3. Results

### 3.1 Adolescent perceptions of a healthy diet

Four themes emerged about adolescent perceptions of a healthy diet. The first theme centred on the idea that healthy diets are composed of meals that include a variety of food groups to provide a range of nutrients and vitamins. One female adolescent explained:

> “Eating well for me means eating all six nutrients… I can tell when there are carbohydrates, lipids, salts, vitamins in the food…. eating rice with fish sauce, orange, for me that’s eating well.” (FG_F, high school, rural, south Benin)

A second theme highlighted an emphasis on food quantity. Most participants perceived healthy diets as those that “provide energy”, emphasising foods like staples (rice, cereals, and cassava). A smaller number of participants also mentioned foods that “provide strength” such as ASF. References to ASF were more common among boys than girls. Third, although mentioned in only a few FGDs, food safety was also perceived by some to be part of a healthy diet. Adolescents noted concerns about processing and preparation, particularly for ASF, which deterred their consumption. Fourth, both boys and girls believed they should eat more F&V than they currently did. Perceived optimal frequency of F&V consumption varied widely, from after each meal to once a day to 2-3 times per week for vegetables.

### 3.2 Food behaviours

Three groupings of food behaviours were discussed by participants: (1) food acquisition, preparation, and allocation at home, (2) consumption at home, and (3) consumption at school. Food acquisition and preparation practices were generally homogenous. They were considered the responsibility of mothers or female caretakers. Both boys and girls reported going or accompanying their mothers on market trips and preparing meals. In contrast, food allocation practices varied widely. Some participants highlighted that young children typically ate first and received more food. Others highlighted that older boys were served first and took the quantities they want while other children waited. Yet others mentioned that boys typically received more food compared to girls because girls ate less than boys.

Reported food consumption at home generally aligned with healthy diet perceptions. The most mentioned foods were grains/roots/tuber, pulses, and other vegetables (**Table 2**). FGDs with older adolescents and those in rural areas placed greater emphasis on ASF and dark green leafy vegetables compared to FGDs with younger adolescents and those in urban areas. Although boys perceived ASF as part of a healthy diet more commonly than girls, girls mentioned more types of ASF than boys when describing what they consumed at home. However, all ASF groups were mentioned less than other food groups, which was primarily due to high costs and food safety concerns according to FGD participants. For example:

> “Some people who can afford it eat meat and fish every day, but others who can’t afford it, eat soya instead.” (FG_M, high school, urban, north Benin)

**Table 2.**
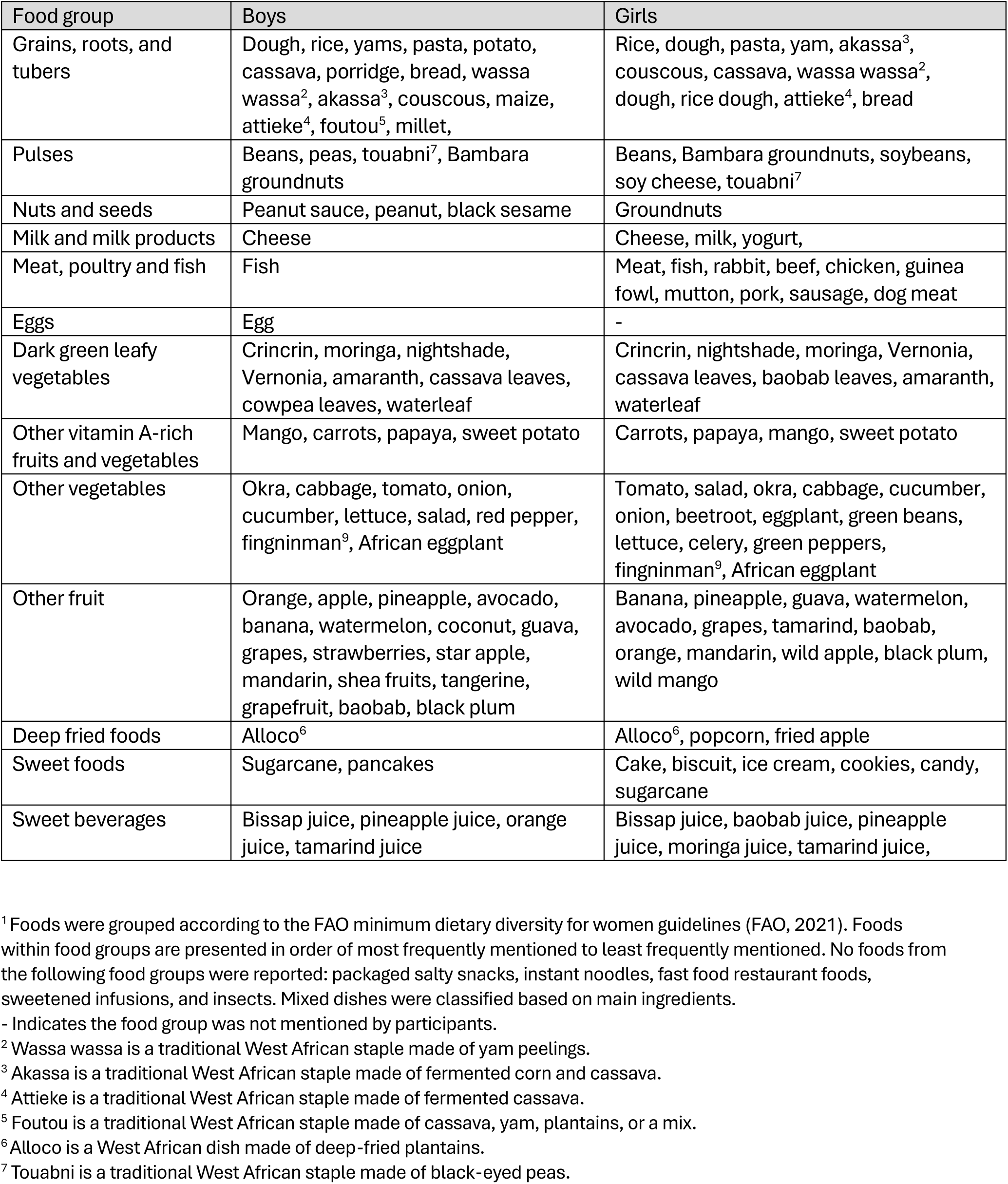
Common foods reported as typically consumed at home by adolescents^1^.

Juice, cheese, and pasta came up more in the south, closer to the capital city of Benin, than in the north. Girls were more likely to mention sweet foods compared to boys. Adolescents shared that they prioritised foods they preferred and foods that were grown in their community, which were more available and affordable:

> “Here, we eat fewer beans because things are getting more expensive and we can’t find money quickly to buy them, and if we grow them, it doesn’t yield much. That’s why we eat fewer beans.” (FG_F, high school, rural, south Benin)

Typical food consumption had changed over time reportedly due to (1) sensitisation at school focused on eating healthy, diverse, and safe foods, (2) changes in food practices at home, with parents preparing and sharing food differently from before (e.g., using individual rather than shared plates), and (3) the effects of climate change on food availability and affordability, specifically indigenous fruits.

The foods adolescents reported typically eating at school varied by gender, age, location, and region. All adolescents mentioned grains/roots/tubers with limited mention of dark green leafy vegetables. Boys mentioned fish, eggs, nuts and seeds, none of which were mentioned by girls. Older adolescents mentioned more diverse foods (e.g., pulses, other fruits, and milk and milk products) than younger adolescents who mentioned primarily grains/roots/tubers and few other fruits. Adolescents in rural schools mentioned more types of foods compared to those in urban schools. In the south, adolescents mentioned juices and sweets (e.g., cookies, biscuits) more compared to adolescents in the north, who typically mentioned traditional staples and pulses.

### 3.3 Drivers of F&V food choice

Three DFC dimensions emerged as key for F&V: intrapersonal drivers, socio-cultural drivers, and FE drivers (**Table 3**). The intrapersonal drivers were (1) knowledge, attitudes, beliefs, and perceptions, (2) preferences, and (3) goals and prioritisation. The social-cultural drivers were family and media influences, peer relationships, cultural values and practices, and religion. The FE drivers were availability, affordability, convenience, and desirability.

**Table 3.** Summary of key findings on adolescent drivers of food choice with respect to fruit and vegetables (F&V)^1^.

| Theme | Definition | Example quote | Respondent | Quote number |
| --- | --- | --- | --- | --- |
| **Intrapersonal drivers** |  |  |  |  |
| Knowledge, attitudes, beliefs, and perceptions | Any mention of perceived health and nutrition benefits of F&V | It depends on one's focus...we are often told that if you eat fruits, you will go 101 years without dying. For me, that's what motivates me to eat fruit, because I don't want to die soon. | FG_F, middle school, rural, north Benin | 1 |
|  |  | Vegetables often strengthen white blood cells, which provide protection against the proliferation of microbes. Those who don't consume enough will quickly catch illnesses. | FG_M, middle school, urban, north Benin | 2 |
|  |  | I also like bitter sauce (*sauce amer*), because it helps to release waste from the body and I like crin crin because it fights malaria.<sup>2</sup> | FG_F, middle school, rural, north Benin | 3 |
|  |  | Fruits allow our body to renew cells in order to digest waste and toxic elements that are harmful to our body. | FG_M, high school, urban, north Benin | 4 |
|  |  | Lemon too, if you're a bit overweight and want to lose a little weight, lemon helps you lose a little weight. | FG_M, middle school, urban, south Benin | 5 |
| Preferences | Any mention of personal preferences (e.g., taste) for specific F&V | I think consuming fruits and vegetables is very good for one's health. I especially like the small, naturally sweet flavors that fruits such as banana, orange, and papaya offer. | FG_F, high school, rural, south Benin | 6 |
|  |  | Here, now, I eat what I prefer to eat every day. But at home, where there is dad and mom, for example, there is a sauce that we children do not eat, though she really likes it with dad. It's called... I don't really know the name in French; in our language we call it 'amanvivè' in Fon (vernonia). It's a vegetable and it is often bitter. When our mother wants to prepare it, she already knows, so she makes two different sauces in the house at that time. She makes one separately for herself and dad, and another one separately for the children. | FG_M, high school, urban, north Benin | 7 |
| Goals and prioritization | Any mention that beauty-related desires influence F&V choices and consumption | I saw that once when they eat vegetables, they speak well. For example, a woman told me that she doesn't like sticky sauce, that she prefers vegetables because it gives a beautiful voice, attractive, and allows you to charm men. | FG_F, high school, urban, north Benin | 8 |
| Socio-cultural drivers |  |  |  |  |
| Family influence | Any mention that family and/or parental preferences influence F&V choices and consumption | That's what my parents do; they are used to eating vegetables, so now I eat them too. | FG_M, high school, urban, north Benin | 9 |
|  |  | Sometimes parents tell us not to eat too much sugary fruit because boys shouldn't eat too much sugary fruit. | FG_M, middle school, rural, south Benin | 10 |
|  |  | Often it's the parents... They often say that some fruits aren't very good and that we shouldn't overindulge in them. | FG_M, middle school, rural, south Benin | 11 |
| Media influence | Any mention that social or traditional media influence F&V choices and consumption | We eat vegetables because, for example, we are in school session right now, and even on television we see that vegetables are good for our health, so that's what encourages us to eat vegetables. | FG_M, high school, urban, north Benin | 12 |
| Peer relationships | Any mention that peer relationships influence F&V choices and consumption | Yes, I think it influences our eating habits. For example, when I see one of my friends eating something I like, I might take out the money and buy it myself too. | FG_F, middle school, rural, south Benin | 13 |
| Cultural values and practices | Any mention that cultural values and practices influence F&V choices and consumption | When we eat bitter vegetables with the paste, our classmates often make fun of us. Because, in reality, up north, they don't consider things like that to be food. | FG_M, middle school, urban, north Benin | 14 |
| Religion | Any mention that religion influences F&V choices and consumption | When we consume moringa, for example, others say that moringa is not for everyone, that it's mainly Muslims who eat moringa because they often make *dambou* with it. | FG_M, middle school, urban, north Benin | 15 |
| Food environment |  |  |  |  |
| Availability | Any mention that availability at home, in the community, or in markets influences F&V choices and consumption | In my opinion, if it's vegetables, I eat them every evening because they are available during the rainy season, and during the dry season, people grow them in gardens. But fruits ... well, there's a season, and during that season, you can find those fruits. Currently, we are in the dry season, so we can't find mangoes, but we can find papayas. | FG_F, high school, rural, north Benin | 16 |
|  |  | Because at home, the fruits don't grow quickly. It has seasons. If the fruit is in season, we eat properly. But if it's not [in season], then it's rare. | FG_F, middle school, rural, south Benin | 17 |
|  |  | These foods are not consumed because they are difficult to grow. And also, when you do grow them, and you don't find much [of it], so you have to sell them at a high price to recoup what you put into growing them. | FG_F, high school, rural, north Benin | 18 |
|  |  | There are also climate changes, for example, this year there are no mangoes that have come out, normally they should have come out, but because of the climate the mangoes did not produce this year. | FG_M, high school, rural, south Benin | 19 |
| Affordability | Any mention that affordability of F&V influences F&V choices and consumption | It is because of a lack of financial resources that we no longer consume juices, fruits, and vegetables today. It is due to the lack of financial means. | FG_M, high school, urban, south Benin | 20 |
|  |  | To prepare vegetables, you have to buy a large black nightshade for five hundred francs, and even then, you won't get enough to eat twice. For the five hundred-franc black nightshade, you need ginger, which is expensive. Everything you buy to make the sauce ready and good to eat, you have to spend about two thousand francs before the pot is even good. All of this means we lack the means, and we can't eat it properly. But during the rainy season, we have it growing in our fields, in front of our houses. We can eat it; because you already have your vegetables, you can get some sesame seeds, and there's also some tomato. If you chop some onion, it's already good. | FG_F, middle school, urban, north Benin | 21 |
|  |  | If we don't consume them, it's because we don't have the means; otherwise, if you go to the market, you will find them. | FG_F, middle school, urban, north Benin | 22 |
|  |  | We want to, but there's no money. If you have money, watermelon is available here. We do it a lot, but it's money that we lack. | FG_F, middle school, urban, north Benin | 23 |
| Convenience | Any mention that convenience of F&V influences F&V choices and consumption | For example, with sugarcane, you first have to remove the peel before eating it; that's why I don't eat it. Also, you can't eat mango in town; people will look at you and make fun of you, whereas if I'm at home, I feel comfortable. Besides, when you eat mango during the day, flies bother you. That's why I eat it in the evening. That's why I don't eat fruits frequently. | FG_M, middle school, urban, south Benin | 24 |
| Desirability | Any mention that desirability of F&V influences F&V choices and consumption | Nowadays, some vegetables are poorly washed; if we eat them, we get sick. Also, the fertilizers that are used make us sick as well. They contain acid that makes us ill. That is why many people say that some vegetables are not good to eat. | FG_M, middle school, urban, south Benin | 25 |
|  |  | We also had an NGO that came to the school as part of the "let's consume local" initiative. They also told us that everything that comes from outside is not good. They said that pesticides are used and that products that are not good for the body are often applied in the fields. | FG_M, middle school, urban, north Benin | 26 |

<sup>1</sup> Abbreviations used: FG\_F, focus group with female adolescents; FG\_M, focus group with male adolescents; F&V, fruits and vegetables.
<sup>2</sup> Crin crin is a dark green leafy vegetable, indigenous to West Africa. In Benin, it is usually prepared as a slimy sauce served with staples like fufu or pounded yams.

#### 3.3.1 Intrapersonal drivers

The most prominent intrapersonal driver was adolescents’ knowledge, attitudes, beliefs, and perceptions about F&V’s health and nutrition benefits. Perceived benefits included: prevention of disease (e.g., malnutrition, anaemia, and malaria); provision of healthy nutrients, vitamins, and energy; supporting a long life; promoting digestive health; and detoxification (Table 3, Quotes 1-4). Across all FGDs, adolescents perceived that certain fruits have specific health benefits. For example, papaya and oranges were perceived as good for digestion, watermelon for blood/iron, and lemons for weight loss (Table 3, Quote 5).

Taste preferences were also reported to influence F&V consumption. For example, adolescents reported liking the sweetness of fruits and the slimy and sticky texture of certain vegetables (Table 3, Quote 6). In contrast, disliking the taste of certain F&Vs deterred adolescents’ consumption (Table 3, Quote 7). Lastly, beauty-related goals and prioritisation among girls motivated them to eat F&V. As one participant explained, consuming F&Vs contributes to clear skin, beauty, charm, and a strong voice (Table 3, Quote 8).

#### 3.3.2 Socio-cultural drivers

Family and media influences, peer relationships, cultural values and practices, and religion were the most salient socio-cultural drivers of F&V consumption that emerged, particularly among boys (Table 3, Quotes 9-15). Family influence was both a facilitator and a barrier to F&V consumption (Table 3, Quotes 9-11). Adolescents explained that parents’ limited knowledge about F&V benefits and religious beliefs resulted in reduced purchasing of F&V, hindering adolescents’ consumption. Media influences through television facilitated F&V consumption (Table 3, Quote 12). Peer relationships emerged as a facilitator, with adolescents reporting to model peer behaviour in food choice and specifically to eat F&V because their peers did (Table 3, Quote 13). Cultural and religious values and practices, however, reflected food-related cultural stigma, where certain foods were perceived to be for specific cultural groups (Table 3, Quotes 14-15). Adolescents were also sometimes teased for eating these foods.

#### 3.3.3 FE

Availability, affordability, convenience, and desirability emerged as prominent barriers to F&V consumption with no differences across adolescent gender, age, location, or region (Table 3, Quotes 16-26). Regarding availability, adolescents explained that F&V were available in markets throughout the year, with some seasonal fluctuations. Vegetables were often grown in home gardens, which increased their availability and consumption (Table 3, Quotes 16). Fruits were reportedly consumed based on seasonal availability (Table 3, Quotes 17). In addition, adolescents reported that when F&Vs that were less available in the community were grown by households, these F&Vs were often prioritized for sale rather than self-consumption so that families could benefit from income from the sale (Table 3, Quote 18). Lastly, climate shocks like limited rain and increased droughts reportedly reduced F&V availability from prior years (Table 3, Quote 19).

Low affordability (i.e., high prices and lack of financial resources) was another frequently reported barrier to F&V consumption (Table 3, Quotes 20-23). For vegetables, the cost of preparation in the preferred manner also hindered consumption (Table 3, Quote 21). Regarding convenience, adolescents reported avoiding fruits that required peeling or were not easily edible such as mangoes, reflecting the importance of convenience (Table 3, Quotes 24). The only desirability barrier mentioned was lack of trust in F&V safety (Table 3, Quotes 25-26).

### 3.4 Free listing and pile sorting

The findings from the free-listing and pile sorting exercises for facilitators and barriers to F&V consumption aligned with FGD findings. During the free listing, adolescents identified health goals and prioritisation and desirability as the main facilitators to F&V consumption (**Figure 2**) and availability and affordability as the main barriers to F&V consumption (**Figure 3**). We observed some variation by region, location, and age. Younger adolescents and those in the south mentioned goals and prioritisation as a facilitator of F&V consumption more times than older adolescents and those in the north, respectively (**Supplemental Figures 1-2**). Desirability was mentioned more times in FGDs with older adolescents than younger adolescents. Affordability was reported more prominently as a barrier to fruit consumption in the north than in the south (**Supplemental Figure 3**). Availability was a more prominent barrier to vegetable consumption among younger than older adolescents and a more prominent barrier to F&V consumption in rural than urban schools (**Supplemental Figure 4**).

**Figure 2.**
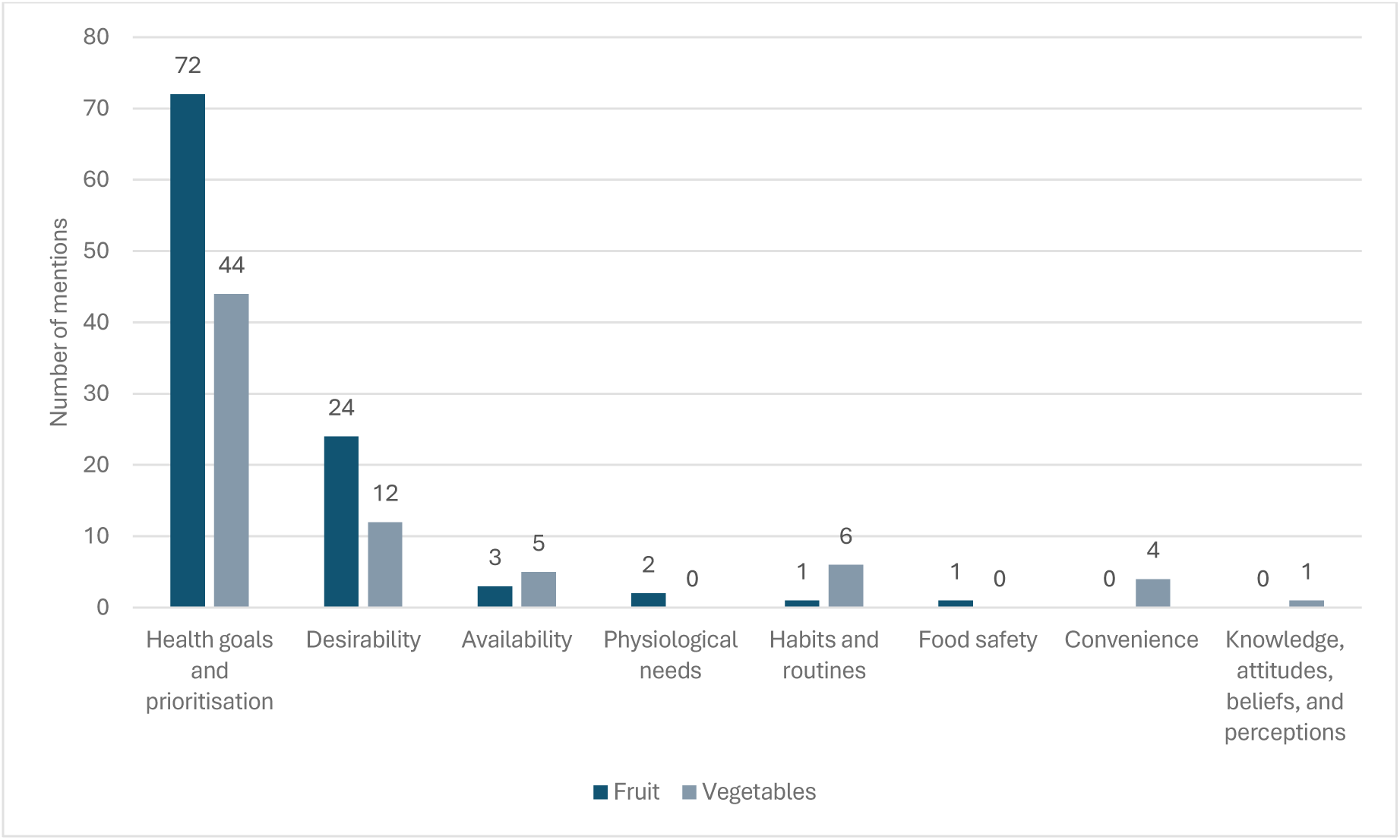
Facilitators of fruit and vegetable consumption among adolescents

**Figure 3.**
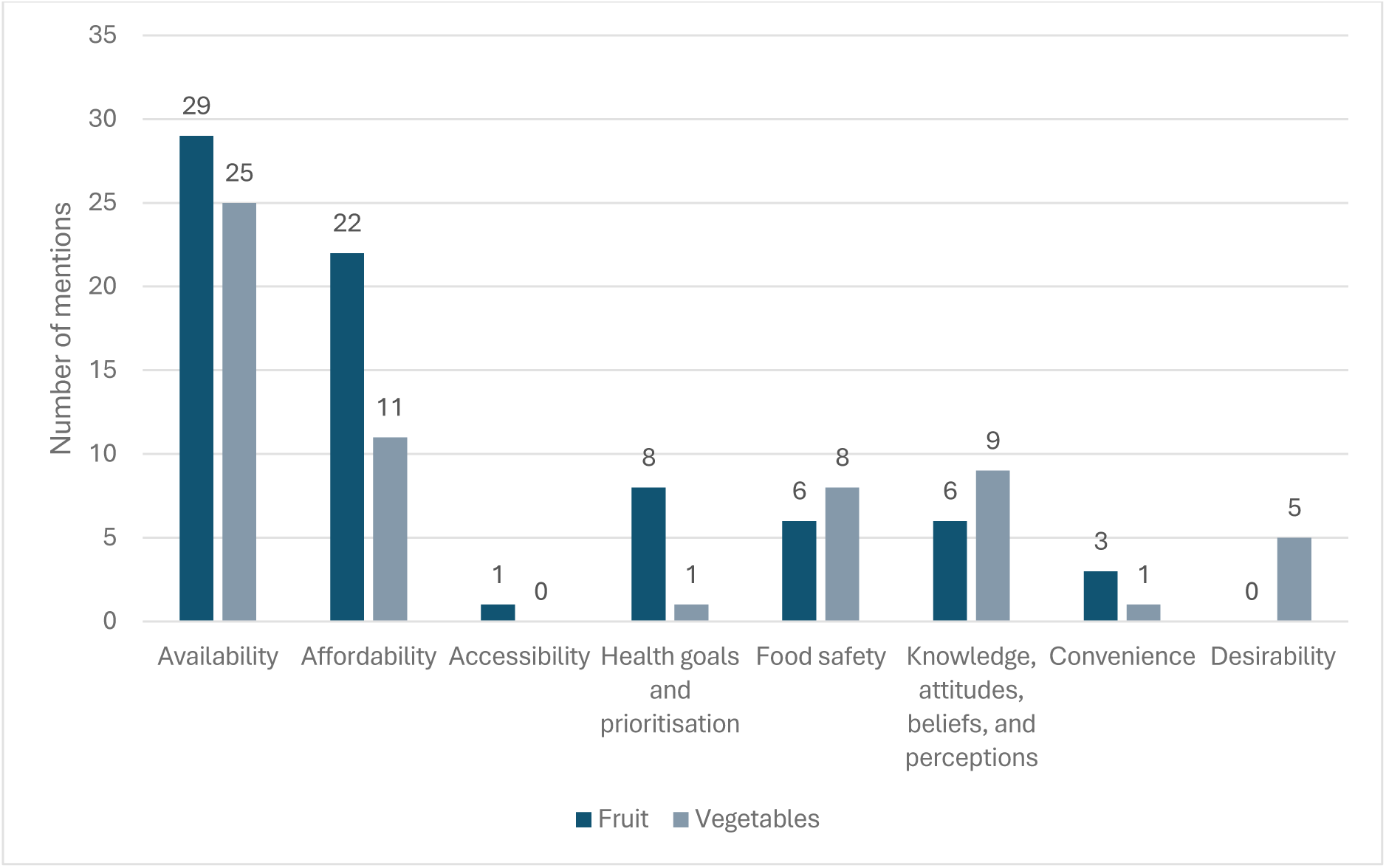
Barriers to fruit and vegetable consumption among adolescents

The pile sorting exercise confirmed these results. Health goals and prioritisation, desirability, and availability were the most important facilitators for F&V consumption, while convenience was also among the most important facilitators for vegetable consumption. The most important barriers to F&V consumption were affordability, availability, and food safety.

During the free listing, adolescents identified several fruits (n=13) and vegetables (n=22) as undesirable or prohibited, citing perceived health risks, personal, cultural, and religious beliefs. These perceptions reduced the desirability of foods such as okra, amaranth, mango, and lemon.

### 3.5 School SSOs

The SSOs were designed to understand food availability at schools. All five schools observed had vendors inside the school and three schools had vendors outside the school within a 100m radius. The average number of vendors within schools was 9 (range 1-20) and outside of schools was 3 (range 0-6). More food groups were available among vendors inside than outside the schools (**Table 4**). Notably, bananas were the only fruit available, observed at both urban schools but none of the three rural schools. During the SSOs, vendors explained that they chose what to sell based on their own cultural preferences or personal experiences, student demand or appreciation, and the input costs of preparing certain items or dishes. During the validation workshop, vendors explained that fruits did not make students feel full and they spoiled quickly; therefore, they did not prioritise selling them.

**Table 4.** Foods available within and outside of schools based on semi-structured observations in five schools^1^.

|  | Number of schools where the food group was available |  |
| --- | --- | --- |
|  | Inside the school | Outside the school<sup>2</sup> |
| Number of schools with type of vendor | 5 | 3 |
| Grains, roots, and tubers | 4 | 3 |
| Pulses | 3 | 0 |
| Nuts and seeds | 2 | 1 |
| Milk and milk products | 3 | 2 |
| Meat, poultry and fish | 4 | 3 |
| Eggs | 3 | 2 |
| Dark green leafy vegetables | 2 | 1 |
| Other vitamin A-rich fruits and vegetables | 2<sup>†</sup> | 1<sup>†</sup> |
| Other vegetables | 4 | 3 |
| Other fruit | 2 | 0 |
| Deep fried foods and packaged salty snacks | 5 | 3 |
| Sweet foods | 5 | 2 |
| Sweet beverages | 3 | 3 |
| Packaged water (bottle or sachet) | 4 | 3 |

<sup>1</sup> Foods were grouped according to the FAO minimum dietary diversity for women guidelines (FAO, 2021). No foods from the following food groups were reported: instant noodles, fast food restaurant foods, sweetened infusions, and insects. Mixed dishes were classified based on main ingredients. Vegetables were primarily available as sauces to accompany staples.
<sup>2</sup> Food outlets within 100 meters outside the school that were open at the time of the observation were counted.
<sup>†</sup> Only other vitamin A-rich vegetables were available on the day of the observation.
<sup>‡</sup> Banana was the only fruit available at the time of the observation.
<sup>\*</sup> Only fish was available at the time of the observation.

## 4. Discussion

This qualitative study used data from 16 FGDs with 126 participants and five SSOs to explore factors that influence adolescent F&V food choices. We drew on data from two regions, younger and older adolescents, boys and girls, and urban and rural areas. We identified four main drivers of food choice for adolescent F&V consumption: intrapersonal (awareness and knowledge of healthy eating and the nutrition and health benefits of F&V, taste preferences, and health prioritisation), socio-cultural (family and peer influences), and FE (low availability low affordability, low convenience, and low desirability due to food safety concerns).

Consistent with prior research, adolescent DFC are a dynamic interplay of multiple dimensions (Mukanu et al., 2022; Neufeld et al., 2022; Verstraeten et al., 2016). Similar to prior studies in urban Benin and among girls (Mama Chabi et al., 2022; Nago et al., 2012), we found that limited availability, low affordability, and food safety concerns persisted as major barriers to F&V consumption. We build on this work by showing these factors hinder F&V consumption regardless of adolescent age, gender, and location. Contrary to this prior research, we found limited evidence that low accessibility influenced adolescent F&V DFC – it emerged as a barrier only in the free listing and pile sorting exercises, without mention in the FGDs. This may be because prior studies in Benin were conducted 5 to 15 years before ours and low accessibility may be less of a barrier today.

Knowledge, attitudes, beliefs, and perceptions were an important adolescent DFC across both genders and age groups. Adolescents generally had positive attitudes toward healthy diets, which they described as those comprising foods that provided the desired energy, strength, and health. Evidence from other traditional FEs like Benin’s shows that adolescents make links between eating well and healthiness (Neufeld et al., 2022). F&V knowledge, attitudes, beliefs, and perceptions were aligned with what adolescents described eating at home and at school, with few exceptions like fruits and ASF. Although adolescents expressed preferences for certain fruits and ASF and perceived benefits from their consumption, reported consumption was low due to high costs, lack of family knowledge that these are healthy foods, and food safety concerns. The strongest gender differences we observed were regarding ASF, where boys mentioned them more than girls as foods that make up a healthy diet, but girls mentioned more types of foods they consumed than did boys. We did not, however, measure adolescent dietary intake and lacked data on the quantity, quality, frequency, and source of food intake and therefore, could not confirm if and how reported knowledge, attitudes, beliefs, and perceptions translate to actual food consumption. Nevertheless, positive attitudes toward nutrition and healthy eating have been linked to better diet quality in West African adolescents (Klemm et al., 2025). Notably, some knowledge and beliefs were incorrect (e.g., watermelon being good for iron/blood) or did not align with local recommendations (e.g., perceived optimal F&V consumption frequency) (FAO, 2015). Other studies from urban Benin have documented poor dietary knowledge, but adequate dietary attitudes and practices among school-going children (Bello et al., 2024), supporting a further need for nutrition education.

Consistent with the literature (Mama Chabi et al., 2022; Mukanu et al., 2022; Neufeld et al., 2022), we found that family and peers influenced adolescent F&V choices and behaviours. Our results showed that adolescents actively participated in household food acquisition and preparation. For vegetables, adolescents emulated parental practices and choices. For fruits, however, parental practices were a barrier – if parents did not purchase fruits for the household, adolescents lacked access despite expressed preferences for certain fruits. This finding aligns with evidence from other traditional FEs that family priorities often superseded adolescents’ individual considerations and agency (Neufeld et al., 2022). In addition to family influences, peer influences shaped F&V choices and acceptability, which was not surprising given that social acceptability among peers is a well-documented adolescent DFC (Mukanu et al., 2022; Patton et al., 2016; Ragelienė & Grønhøj, 2020). Unlike prior work in Benin that documented peer pressure for unhealthy food choices (Mama Chabi et al., 2022), in our study, adolescents reported copying their peers to consume more F&V. This difference may be due to differences in study objectives, areas, and target populations. Nevertheless, these findings suggest that interventions to improve adolescent diets should actively engage adolescent family members and peers both in the design and implementation phases, and that comprehensive sensitisation at the school, family, and community level is needed.

Further, our findings also showed that the FE was an important DFC. Adolescents were acutely aware, even at their young age, that F&V can be costly and that F&V affordability and availability are influenced by seasonality and climate shocks. Increasingly, adolescents in many African settings recognise the effects of climate change on food access and associated trade-offs when food availability is low (Hall et al., 2019). In addition, availability at school influenced adolescent F&V DFC. Similar to many other low- and middle-income countries where school FEs are increasingly providing unhealthy, cheap, ultra-processed foods (Faber et al., 2019; Mukanu et al., 2022), adolescents in our study had access to multiple unhealthy options such as deep fried and packaged salty snacks, sweets, and sugar-sweetened beverages. Although many schools had foods from healthy food groups too, fruits were notably absent, which vendors explained was because they spoiled quickly and did not make adolescents full. These results show that healthy and unhealthy options co-exist in the school FE. Interventions to improve adolescent diets should consider activities to support adolescents’ choices of healthier options when presented with both. Interventions that improve vendors’ fruit storage could help improve fruit accessibility at school, and consequently adolescent fruit consumption considering their expressed preference for certain fruits.

Lastly, we found that taste and texture preferences supported adolescent F&V consumption. However, food safety concerns (e.g., overuse of pesticides during production, handling of ASF by vendors) reduced the desirability of F&Vs and ASF, both nutrient-rich foods that adolescents need for optimal health, nutrition, and development. Other studies in urban Benin have documented food safety as a challenge at schools. In one study, adolescents reported only snacking (e.g., fried foods, sweets) because of food safety concerns: foods not well-covered, lack of school policies on food safety, and vendors not being dressed in a tidy way (Mama Chabi et al., 2022). In an earlier study, adolescents reported not eating F&V at school because of how they were handled and prepared (Nago et al., 2012). Together, these findings suggest that adolescents minimise consumption of nutrient-rich foods in response to food safety concerns, which outweigh considerations of health and nutritional benefits, consistent with other settings (Verstraeten et al., 2016). More research to unpack adolescent food safety perception and actual vendor and production practices can help inform future interventions to address food safety concerns.

This study used a large diverse sample with variation by age, gender, location, and region. The high number of FGDs and participants was sufficient to reach saturation and explore variation analytically. Nevertheless, the use of FGDs implies limitations. It is possible that not all adolescents participated fully in the discussion and results reflect dominant opinions and/or views of more vocal participants (Krueger & Casey, 2009). This concern was mitigated by rigorous training on techniques to establish a comfortable and friendly environment for discussion and encouraging all participants to speak. Homogenising the FGDs in terms of age and gender also likely helped mitigate dominant voices (Smithson, 2000). Additionally, moderator bias cannot be fully ruled out, despite rigorous training and the use of standardised topic guides with neutral probing questions.

In conclusion, this qualitative study showed that a dynamic interplay of intrapersonal, socio-cultural, and FE factors influences adolescent F&V food choices in Benin. Although adolescent knowledge, motivation, and positive peer influences facilitated F&V consumption, they were insufficient when family influences, availability, affordability, and food safety were unsupportive. Approaches to increase F&V consumption and improve adolescent diets should address the intrapersonal, socio-cultural and FE dimensions. Comprehensive actions to influence and support adolescent healthy food choices should address affordability and availability constraints and involve both families and peers. Addressing food safety concerns – through improved production and post-harvest practices or policies for school vendors for example – can help build consumer trust and increase nutrient-rich food consumption. Interventions that actively and meaningfully engage adolescents in their design and implementation are more likely to be effective and responsive to adolescent diet, health, and nutrition needs (Baird et al., 2025).

## Data Availability

De-identified transcripts, the study codebook, and topic guides are available upon reasonable request from the corresponding author.

## Acknowledgements

We would like to thank respondents for their time and participation in the study. We are grateful to Chèrif Issifou for his support in analysing results from the pile sorting and free listing, and for consolidating findings from the community validation workshop. We acknowledge Ahmed Dougouna Djoufouna for his support in analysing the structured observations.

## Author contributions

ADD, LB, and EI designed the study. ADD, SB, AP, and IMM oversaw enumerator training. ADD, SB, AP, and IMM supervised data collection. ADD, SE, and EI conducted the data analysis. ADD, LB, and EI drafted the manuscript. All authors reviewed and approved the manuscript.

## References

Afshin, A., Sur, P. J., Fay, K. A., Cornaby, L., Ferrara, G., Salama, J. S., Mullany, E. C., Abate, K. H., Abbafati, C., Abebe, Z., Afarideh, M., Aggarwal, A., Agrawal, S., Akinyemiju, T., Alahdab, F., Bacha, U., Bachman, V. F., Badali, H., Badawi, A., … Murray, C. J. L. (2019). Health effects of dietary risks in 195 countries, 1990–2017: A systematic analysis for the Global Burden of Disease Study 2017. The Lancet, 393(10184), 1958–1972. 10.1016/S0140-6736(19)30041-8

Baird, S., Choonara, S., Azzopardi, P. S., Banati, P., Bessant, J., Biermann, O., Capon, A., Claeson, M., Collins, P. Y., De Wet-Billings, N., Dogra, S., Dong, Y., Francis, K. L., Gebrekristos, L. T., Groves, A. K., Hay, S. I., Imbago-Jácome, D., Jenkins, A. P., Kabiru, C. W., … Viner, R. M. (2025). A call to action: The second Lancet Commission on adolescent health and wellbeing. The Lancet, 405(10493), 1945–2022. 10.1016/S0140-6736(25)00503-3

Bello, F., Koukou, E., Bodjrenou, S., Termote, C., Azokpota, P., & Hounkpatin, W. A. (2024). Food and nutrition knowledge, attitudes and practices among children in public primary school with canteens in southern Benin: A case study. BMC Nutrition, 10(1), 40. 10.1186/s40795-024-00857-7

Biswas, T., Townsend, N., Huda, M. M., Maravilla, J., Begum, T., Pervin, S., Ghosh, A., Mahumud, R. A., Islam, S., Anwar, N., Rifhat, R., Munir, K., Gupta, R. D., Renzaho, A. M. N., Khusun, H., Wiradnyani, L. A. A., Radel, T., Baxter, J., Rawal, L. B., … Mamun, A. (2022). Prevalence of multiple non-communicable diseases risk factors among adolescents in 140 countries: A population-based study. eClinicalMedicine, 52, 101591. 10.1016/j.eclinm.2022.101591

Blake, C. E., Frongillo, E. A., Warren, A. M., Constantinides, S. V., Rampalli, K. K., & Bhandari, S. (2021). Elaborating the science of food choice for rapidly changing food systems in low-and middle-income countries. Global Food Security, 28, 100503. 10.1016/j.gfs.2021.100503

Bliznashka, L., Pather, K., Mitchodigni, I. M., Hess, S. Y., & Olney, D. K. (2024). Diets, fruit and vegetables consumption, and nutritional status in Benin: A scoping review. *Maternal & Child Nutrition*, e13747. 10.1111/mcn.13747

Bodjrènou, S., Koukou, E., Termote, C., Mitchodigni, I., Joyce, C., Gelli, A., Olney, D. K., & Honeycutt, S. (2023). Food and nutrient intake among children in rural Benin. https://hdl.handle.net/10568/137409

Boncyk, M. (2023, March 14). Drivers of food choice and food choice behaviors constructs. Drivers of food choice for food system transformation in South Asia, Dhaka.

Delisle, H., Ntandou-Bouzitou, G., Agueh, V., Sodjinou, R., & Fayomi, B. (2012). Urbanisation, nutrition transition and cardiometabolic risk: The Benin study. British Journal of Nutrition, 107(10), 1534–1544. 10.1017/S0007114511004661

Faber, M., De Villiers, A., Hill, J., Van Jaarsveld, P. J., Okeyo, A. P., & Seekoe, E. (2019). Nutrient profile and energy cost of food sold by informal food vendors to learners in primary and secondary schools in the Eastern Cape, South Africa. Public Health Nutrition, 22(3), 521–530. 10.1017/S1368980018003464

FAO. (2015). Food-based dietary guidelines—Benin. http://www.fao.org/nutrition/education-nutritionnelle/food-dietary-guidelines/regions/benin/fr/

FAO. (2021). Minimum dietary diversity for women. FAO. 10.4060/cb3434en

Guo, Y., Azazh, A., Kouchica, C. E. B., Gao, S., Han, X., Chang, J., Yu, P., Fan, Y., & Wang, M. (2026). Non-communicable diseases among adolescent and young adult females in sub-Saharan Africa. International Journal of Epidemiology, 55(2), dyag022. 10.1093/ije/dyag022

Hall, B. J., Garabiles, M. R., De Hoop, J., Pereira, A., Prencipe, L., & Palermo, T. M. (2019). Perspectives of adolescent and young adults on poverty-related stressors: A qualitative study in Ghana, Malawi and Tanzania. BMJ Open, 9(10), e027047. 10.1136/bmjopen-2018-027047

Hennink, M., & Kaiser, B. N. (2022). Sample sizes for saturation in qualitative research: A systematic review of empirical tests. Social Science & Medicine, 292, 114523. 10.1016/j.socscimed.2021.114523

IHME. (2015). GBD Compare. IHME, University of Washington. Institute for Health Metrics and Evaluation. https://vizhub.healthdata.org/gbd-compare

IHME. (2026). Benin. https://www.healthdata.org/research-analysis/health-by-location/profiles/benin

INSAE & ICF. (2019). Enquête Démographique et de Santé au Bénin, 2017-2018. INSAE et ICF. https://dhsprogram.com/publications/publication-FR350-DHS-Final-Reports.cfm

Karanja, A., Ickowitz, A., Stadlmayr, B., & McMullin, S. (2022). Understanding drivers of food choice in low- and middle-income countries: A systematic mapping study. Global Food Security, 32, 100615. 10.1016/j.gfs.2022.100615

Klemm, J., Muli, S., Oluwagbemigun, K., Parlasca, M., Crentsil, A., Ogum, D., Quartey, P., Laar, A., Lartey, A., Borgemeister, C., & Nöthlings, U. (2025). Individual-Level Drivers of Food Choices and Diet Quality Among Adolescents in Urban West Africa: Evidence From Accra, Ghana. Maternal & Child Nutrition, 21(2), e13775. 10.1111/mcn.13775

Krueger, R. A., & Casey, M. A. (2009). Focus Groups: A Practical Guide for Applied Research. Sage Publications.

Mama Chabi, S., Fanou-Fogny, N., Nago Koukoubou, E., Deforche, B., & Van Lippevelde, W. (2022). Factors Explaining Adolescent Girls’ Eating Habits in Urban Benin: A Qualitative Study. Adolescents, 2(2), 205–219. 10.3390/adolescents2020017

Miller, V., Cudhea, F., Singh, G., Micha, R., Shi, P., Zhang, J., Onopa, J., Karageorgou, D., Webb, P., & Mozaffarian, D. (2019). Estimated Global, Regional, and National Cardiovascular Disease Burdens Related to Fruit and Vegetable Consumption: An Analysis from the Global Dietary Database (FS01-01-19). Current Developments in Nutrition, 3, nzz034.FS01-01-19. 10.1093/cdn/nzz034.FS01-01-19

Mukanu, M. M., Thow, A. M., Delobelle, P., & Mchiza, Z. J.-R. (2022). School Food Environment in Urban Zambia: A Qualitative Analysis of Drivers of Adolescent Food Choices and Their Policy Implications. International Journal of Environmental Research and Public Health, 19(12), 7460. 10.3390/ijerph19127460

Nago, E. S., Verstraeten, R., Lachat, C. K., Dossa, R. A., & Kolsteren, P. W. (2012). Food Safety Is a Key Determinant of Fruit and Vegetable Consumption in Urban Beninese Adolescents. Journal of Nutrition Education and Behavior, 44(6), 548–555. 10.1016/j.jneb.2011.06.006

Neufeld, L. M., Andrade, E. B., Ballonoff Suleiman, A., Barker, M., Beal, T., Blum, L. S., Demmler, K. M., Dogra, S., Hardy-Johnson, P., Lahiri, A., Larson, N., Roberto, C. A., Rodríguez-Ramírez, S., Sethi, V., Shamah-Levy, T., Strömmer, S., Tumilowicz, A., Weller, S., & Zou, Z. (2022). Food choice in transition: Adolescent autonomy, agency, and the food environment. The Lancet, 399(10320), 185–197. 10.1016/s0140-6736(21)01687-1

Olney, D. K., Singh, R. G., & Schreinemachers, P. (2021). Fruit and Vegetables for Sustainable Healthy Diets (FRESH). CGIAR System Organization. https://hdl.handle.net/10568/121106

Our World in Data. (2021). How do actual diets compare to the EAT-Lancet diet? [Https://ourworldindata.org/grapher/eat-lancet-diet-comparison?tab=table].

Patton, G. C., Sawyer, S. M., Santelli, J. S., Ross, D. A., Afifi, R., Allen, N. B., Arora, M., Azzopardi, P., Baldwin, W., Bonell, C., Kakuma, R., Kennedy, E., Mahon, J., McGovern, T., Mokdad, A. H., Patel, V., Petroni, S., Reavley, N., Taiwo, K., … Viner, R. M. (2016). Our future: A Lancet commission on adolescent health and wellbeing. The Lancet, 387(10036), 2423–2478. 10.1016/S0140-6736(16)00579-1

PNLMNT. (2011). Rapport final. Enquête globale sur la santé des élève au Bénin en 2009 (p. 58). https://cdn.who.int/media/docs/default-source/ncds/ncd-surveillance/data-reporting/benin/gshs/country-report-gshs-benin-2009.pdf?sfvrsn=9abd0460_2%26download=true

Ragelienė, T., & Grønhøj, A. (2020). The influence of peers′ and siblings′ on children’s and adolescents′ healthy eating behavior. A systematic literature review. Appetite, 148, 104592. 10.1016/j.appet.2020.104592

Smithson, J. (2000). Using and analysing focus groups: Limitations and possibilities. International Journal of Social Research Methodology, 3(2), 103–119. 10.1080/136455700405172

Sodjinou, R., Agueh, V., Fayomi, B., & Delisle, H. (2009). Dietary patterns of urban adults in Benin: Relationship with overall diet quality and socio-demographic characteristics. European Journal of Clinical Nutrition, 63(2), 222–228. 10.1038/sj.ejcn.1602906

Turner, C., Kalamatianou, S., Drewnowski, A., Kulkarni, B., Kinra, S., & Kadiyala, S. (2020). Food Environment Research in Low- and Middle-Income Countries: A Systematic Scoping Review. Advances in Nutrition, 11(2), 387–397. 10.1093/advances/nmz031

Twig, G., Yaniv, G., Levine, H., Leiba, A., Goldberger, N., Derazne, E., Ben-Ami Shor, D., Tzur, D., Afek, A., Shamiss, A., Haklai, Z., & Kark, J. D. (2016). Body-Mass Index in 2.3 Million Adolescents and Cardiovascular Death in Adulthood. New England Journal of Medicine, 374(25), 2430–2440. 10.1056/NEJMoa1503840

Verstraeten, R., Leroy, J. L., Pieniak, Z., Ochoa-Avilès, A., Holdsworth, M., Verbeke, W., Maes, L., & Kolsteren, P. (2016). Individual and Environmental Factors Influencing Adolescents’ Dietary Behavior in Low- and Middle-Income Settings. PLOS ONE, 11(7), e0157744. 10.1371/journal.pone.0157744

WFP. (2026). HungerMap LIVE. https://hungermap.wfp.org/

WHO. (2016). 2016 GSHS Fact Sheet Benin. https://www.who.int/publications/m/item/2016-gshs-fact-sheet-benin

Yip, C. S. C., Chan, W., & Fielding, R. (2019). The Associations of Fruit and Vegetable Intakes with Burden of Diseases: A Systematic Review of Meta-Analyses. Journal of the Academy of Nutrition and Dietetics, 119(3), 464–481. 10.1016/j.jand.2018.11.007

Zheng, M., Park, S. Y., Bolton, K. A., Foong-Fong Chong, M., Grafenauer, S., & Xi, B. (2025). Dietary Intake Trajectories from Early Life and Associated Health Outcomes: A Systematic Review. Advances in Nutrition, 16(11), 100528. 10.1016/j.advnut.2025.100528

Zhou, J., Shan, S., Wu, J., Song, Y., Zhu, L., Li, Q., Zhang, C., Zhu, Y., Sheikh, A., Rahimi, K., Song, P., & Rudan, I. (2026). Global prevalence of hypertension among children and adolescents aged 19 years or younger: An updated systematic review and meta-analysis. The Lancet Child & Adolescent Health, 10(1), 11–21. 10.1016/2352-4642(25)00281-0

