## Supplemental Figures for "Healthy diet perceptions and drivers of fruit and vegetable food choices among adolescents in Benin: a qualitative study"

**Supplemental Figure 1** Facilitators to fruit consumption among adolescents by region, location, and school type

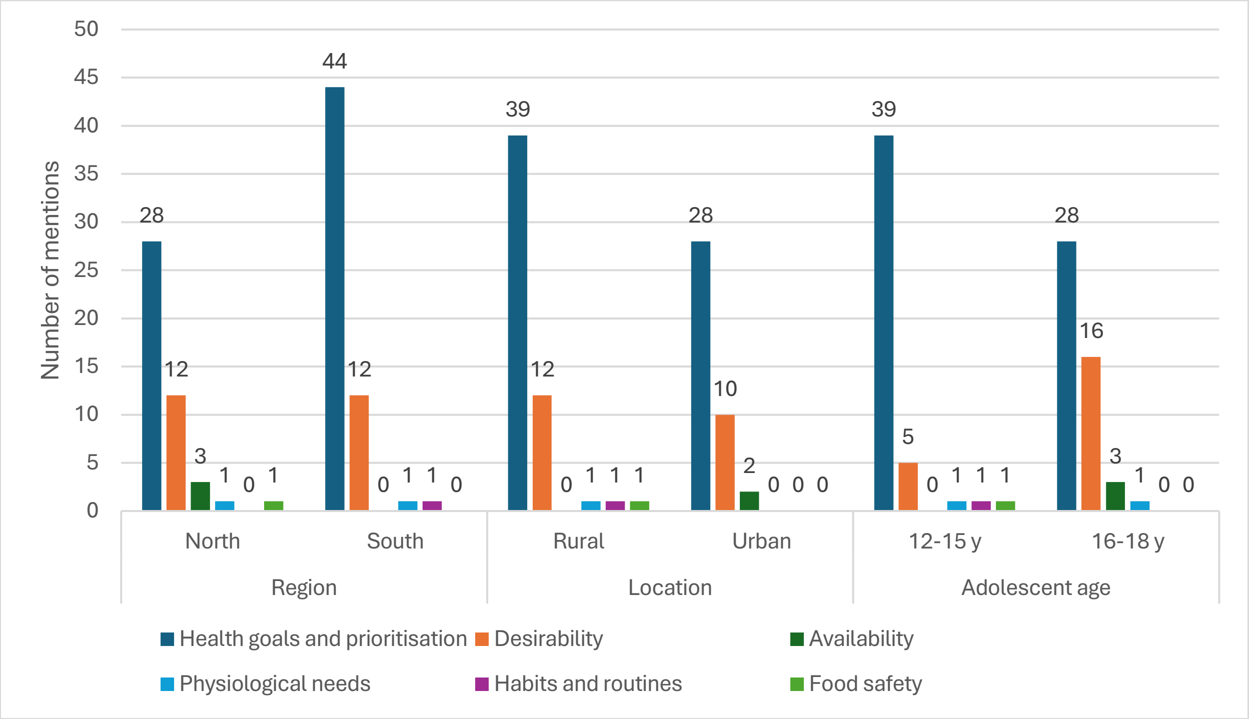


**Supplemental Figure 2** Facilitators consumption among adolescents by region, location, and school type

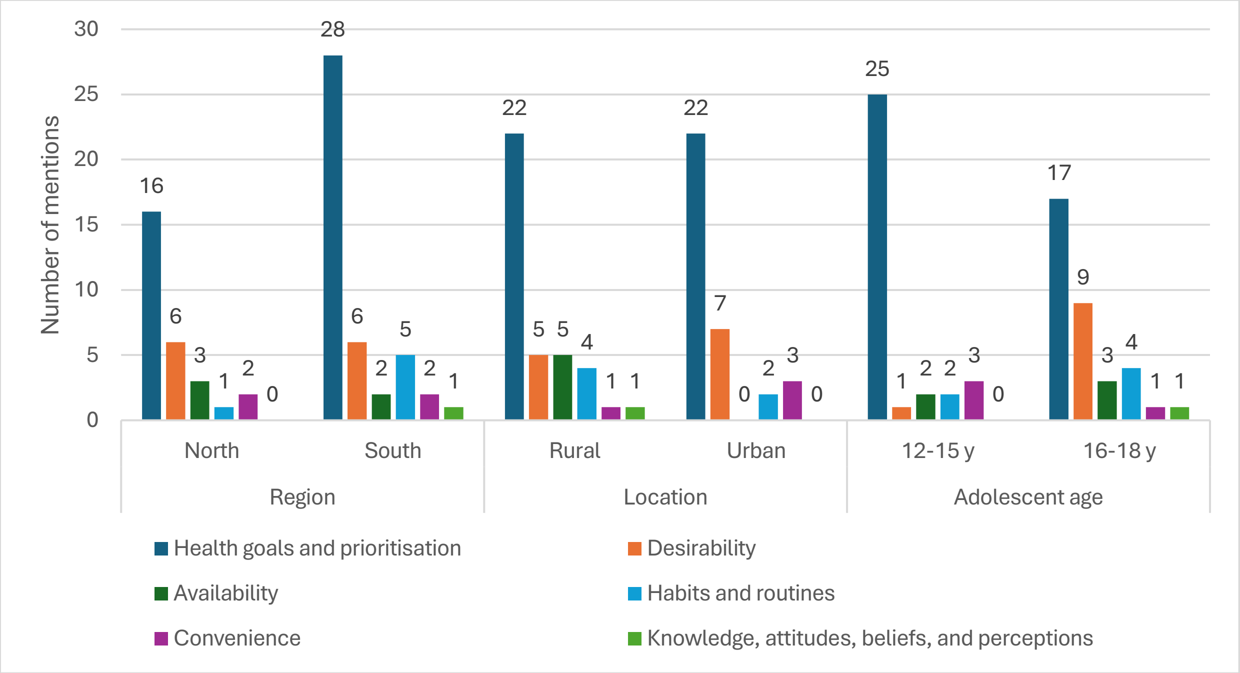

**Supplemental Figure 3** Barriers to fruit consumption among adolescents by region, location, and school type

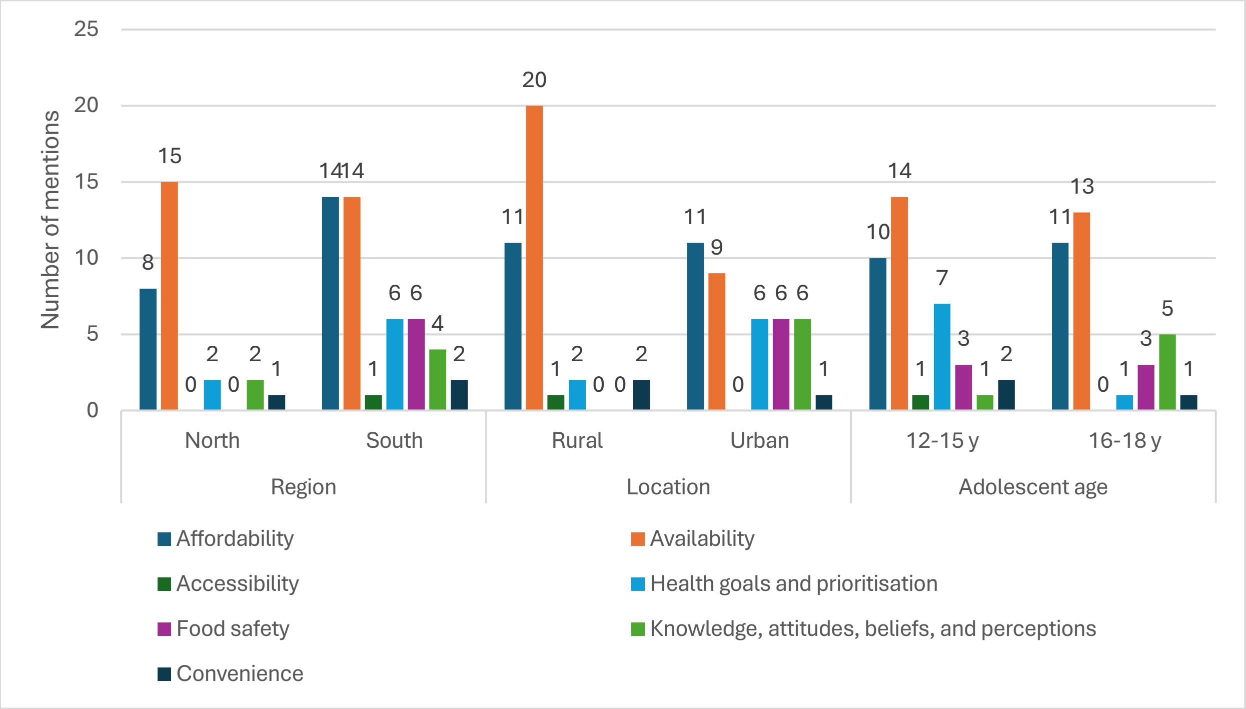


**Supplemental Figure 4** Barriers to vegetable consumption among adolescents by region, location, and school type

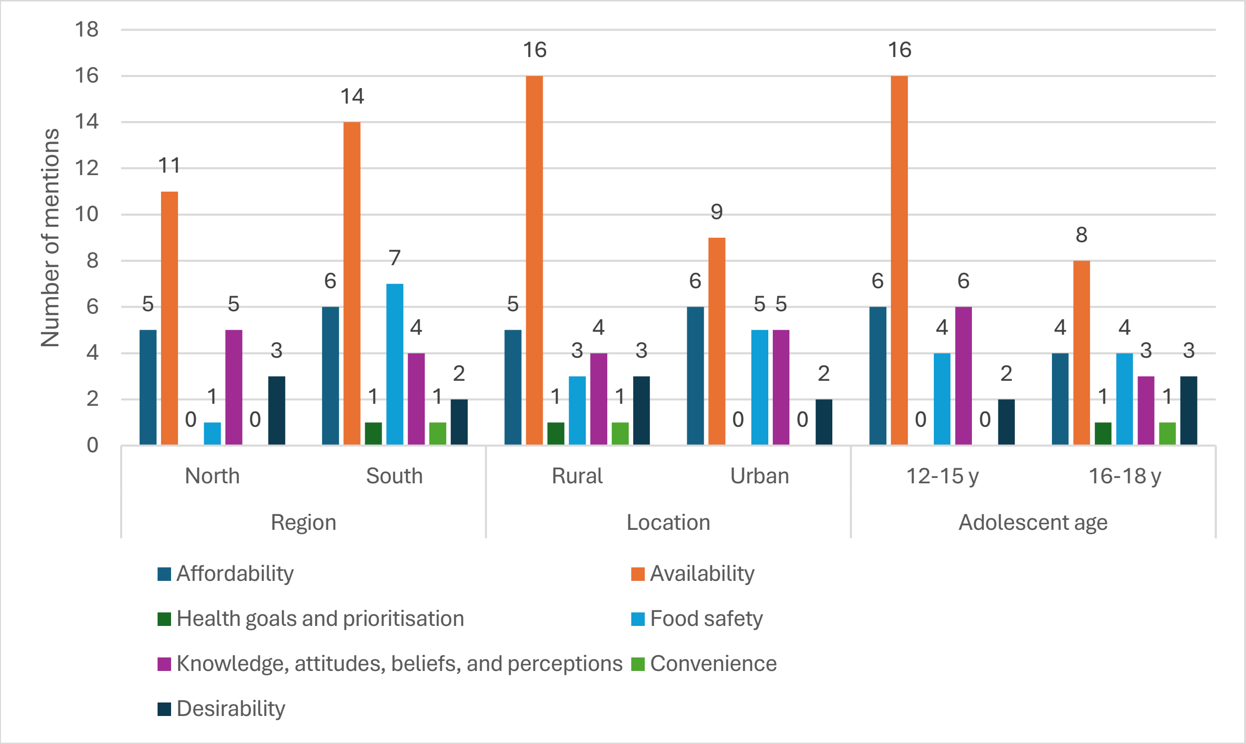
